# Placental fetal vascularization in neonates with congenital heart disease: a pilot retrospective case control study

**DOI:** 10.64898/2026.04.15.26350950

**Authors:** Andrea C Kozai, Takashi Yoshimasu, Maria Chase, Nita R. Chaudhuri, Jai P. Udassi, Bethany Barone Gibbs, Mehdi Hedjazi Moghari

## Abstract

**Background:** Placental function is associated with congenital heart defects (CHD), frequently presenting with malperfusion lesions and small-for-gestational-age size. However, placental villous vasculature in the setting of CHD is understudied. This study evaluated differences in placental, neonatal, and maternal outcomes among maternal/infant dyads with versus without CHD.

**Methods:** We conducted a gestational age- and fetal sex-matched retrospective case control study using specimens prospectively collected by a local biobank. Neonatal outcomes included birthweight, placental weight, and their ratio (placental efficiency). We estimated the proportion of placental villous tissue comprised of fetal vascular endothelial cells (%FVE) using anti-CD34 immunohistochemistry and a pixel count algorithm.

Placental weight multiplied by %FVE estimated the grams of placental tissue comprised of villous vasculature (placental vascular index). Maternal outcomes included hypertensive disorders of pregnancy and gestational diabetes. We compared cases and controls using linear and logistic regression adjusted for maternal smoking and cold ischemia time. Stratified analyses examined associations by preterm birth status.

**Results:** Dyads (n=34 with CHD, n=34 without CHD) had maternal age of 29.4± 4.9 years and were 35.6±4.0 gestational weeks at delivery. Groups had similar placental, neonatal, and maternal parameters. Among preterm neonates, we observed small-to-moderate effect sizes indicating lower placental weight, %FVE, and placental vascular index, and higher placental efficiency, in CHD cases. Among term neonates, moderate effect sizes suggested lower birthweight, placental weight, and placental vascular index in CHD cases.

**Conclusions:** Though differences between groups were not significant, moderate effect sizes suggested that placental vascularization was lower among preterm neonates with CHD.

## Introduction

Congenital heart disease (CHD) is the most common type of birth defect, affecting about 1% of newborns worldwide[1] and approximately 40,000 infants each year in the United States[2]. About one quarter of these infants, especially those with complex CHD, require surgery early in life to survive[3]. CHD exacts a disproportionately high economic burden compared to adult cardiovascular diseases, further underscoring the importance of elucidating mechanisms of disease development[4]. Although some cases of CHD are linked to genetic factors, the exact cause is often unclear and may also involve influences from the maternal environment and placenta during pregnancy[5].

The placenta plays a critical role in supporting fetal development by supplying oxygen and nutrients[6]. In pregnancies complicated by CHD, the placenta is frequently found to have structural and functional abnormalities[7]. These may include poor blood flow from the mother or fetus, areas of placental infarction, abnormal development of placental villi, and smaller placental size[8].

The heart and placenta develop at the same time during early pregnancy and share many biological pathways that control growth and blood vessel formation. Because of this close relationship, disturbances affecting one organ may also affect the other[6,9]. Studies in animal models have shown that disruptions in genes involved in blood vessel development can lead to both fetal heart defects and abnormal placental formation[6,10].

In addition to genetic factors, intracardiac hemodynamic forces such as blood flow are also important[11]. Normal placental blood flow may help support fetal heart development, and reduced flow may limit oxygen and nutrient delivery and compromise heart development. In addition, pregnancies affected by CHD are often associated with complications such as fetal growth restriction, preterm birth, and preeclampsia, which frequently present with placental dysfunction[9]. These outcomes further support the idea that abnormal placentation could play a role in CHD development.

Placentally-mediated adverse pregnancy outcomes such as preeclampsia and preterm birth are associated with long-term maternal cardiovascular consequences[12,13]. Further, preeclampsia and CHD have an intertwined association; a pregnancy complicated by preeclampsia is 38% more likely to also be affected by CHD, and a prior pregnancy complicated by either preeclampsia or CHD is associated with a higher risk of a future pregnancy complicated by the opposite diagnosis[14]. In this way, maternal and fetal outcomes appear to be tightly related to placental development and function.

Prior research has found that newborns with CHD are more likely to have placentas with lower weight and evidence of maternal and fetal malperfusion lesions[7,9,15,16], but precise characterization of placental vascular development in the setting of CHD is limited. Accordingly, the aim of the present study was to compare intravillous placental vascularization, neonatal, and maternal pregnancy outcomes in mother-neonate dyads with versus without CHD. We hypothesized that placental vascularization, weight, and birthweight would be lower among neonates with CHD; we also hypothesized that mothers with a CHD neonate would be more likely to develop a hypertensive disorder of pregnancy.

## Methods

### Study Design and Population

This IRB-approved pilot study employed a retrospective matched case control design. All participants were enrolled in the Magee Obstetric Maternal & Infant (MOMI) Biobank and Database at the Magee Womens Hospital of UPMC, which prospectively enrolls women during pregnancy for tissue bank collection at delivery. Cases were drawn from all available participants who delivered a singleton fetus with a congenital heart defect; we selected 35 cases with at least one of the following diagnoses: transposition of the great arteries; hypoplastic left heart; coarctation of the aorta; tetralogy of Fallot; congenital heart block; severe valve disorders; atrial septal defect; ventricular septal defect; or persistent pulmonary hypertension. Control participants delivered a singleton fetus that did not have a congenital heart defect diagnosis and were matched 1:1 with cases on fetal sex and gestational age at delivery. Participants were excluded if the fetus had a diagnosis of a chromosomal abnormality. All participants consented to use of data and donated tissue through the MOMI Biobank, and the present study was deemed “Not Human Research” by the University of Pittsburgh and West Virginia University ethics boards (STUDY25060063).

### Maternal and Fetal Clinical Data

All clinical data was abstracted from electronic health records by the MOMI Biobank and Database staff and shared on a deidentified basis. Maternal data included age, body mass index at the first prenatal visit, racial and ethnic identity, smoking status, marital status, insurance status, pre-pregnancy hypertension and diabetes status, and pregnancy outcomes including cesarean sections, hypertensive disorders of pregnancy (defined as *de novo* systolic blood pressure ≥ 140 mmHg or diastolic blood pressure ≥ 90 mmHg with or without end-organ involvement, or preeclampsia superimposed on pre-pregnancy chronic hypertension), and gestational diabetes. Fetal data included gestational age at delivery, fetal sex, birthweight, placental weight, and fetal growth (defined as appropriate, small, or large for gestational age using Alexander growth curves). We calculated placental efficiency as the birthweight divided by placental weight.

### Placental Parameters

Placenta tissue was collected following delivery, fixed in formalin, and embedded in paraffin using standardized procedures[17]. The staining, digitization, and pixel-count protocol has been described previously[18]. Briefly, 4µm-thick sections were mounted on slides, deparaffined, stained with anti-CD34 (Leica Biosystems, Buffalo Grove, IL) to highlight fetal vascular endothelial cells, and counterstained with hematoxylin (Sigma-Aldrich, St. Louis, MO) to highlight the remaining placental tissue. The full slide image was digitized at 40x magnification on an Aperio AT2 Whole Slide Scanner. To estimate the proportion of placental tissue occupied by fetal vascular endothelial cells, we centered the image on a grid and selected four 600x600µm boxes (one in each quadrant of the image) that contained only mature intermediate and/or terminal villi (**Figure 1**). The Aperio Positive Pixel Count v9 algorithm (Leica Biosystems, Nussloch, Germany) was used to analyze the number of pixels in each box positively stained for CD34. Finally, we divided the sum of the “positive” and “strong positive” pixels from the four boxes by the sum of the total pixels in the four boxes to find the proportion of pixels representing fetal vascular endothelium (%FVE). In addition to %FVE, we calculated a placental vascular index by multiplying %FVE by the placental weight to estimate the placental weight represented by intermediate and terminal villous vasculature. We used both %FVE and the placental vascular index for analysis.

**Figure 1.**
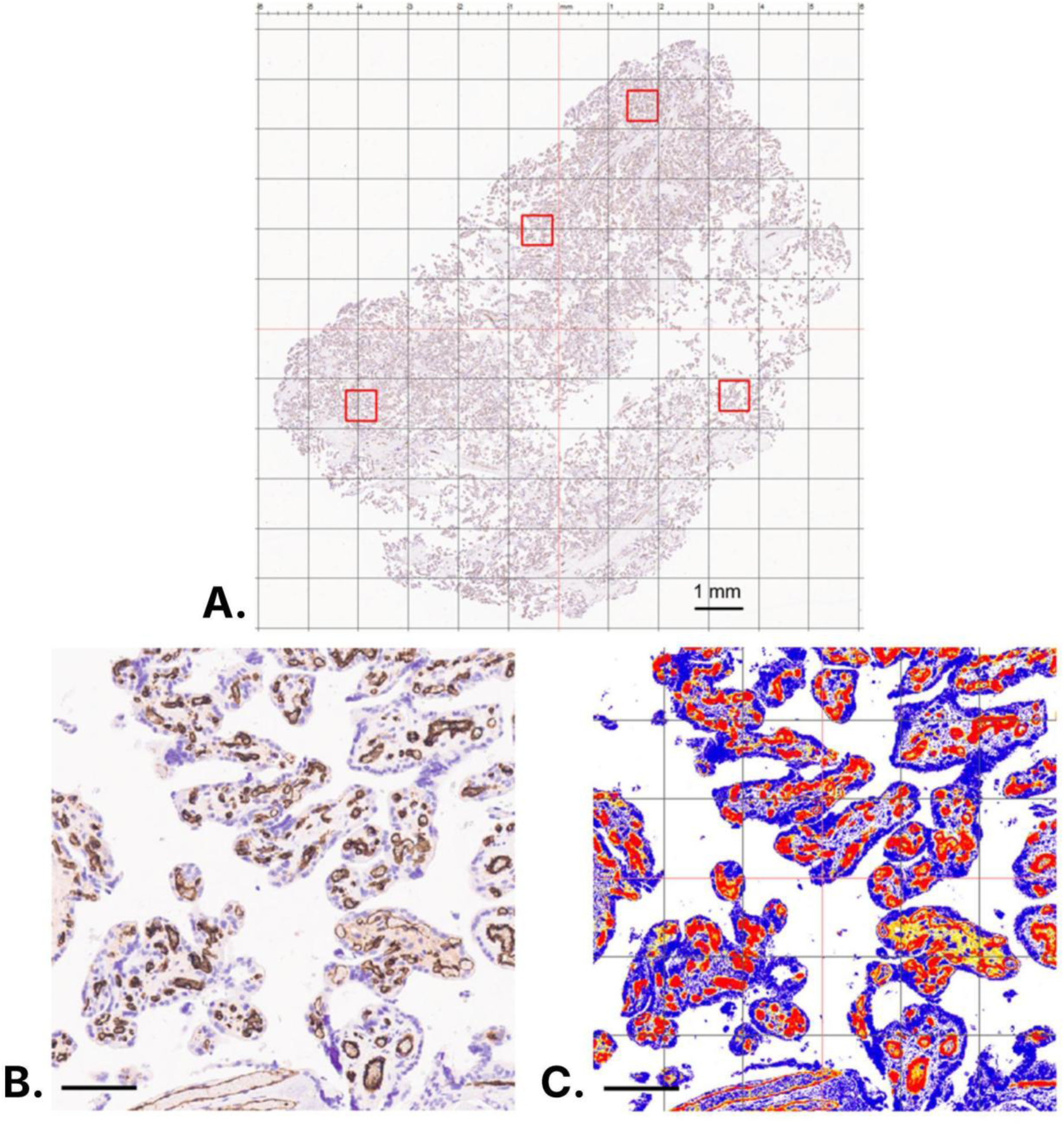
Representative digital images of stained placental tissue for analysis. Scale bars are 100µm unless labeled otherwise. **A.** Whole slide image at 0x magnification centered on a 1mm grid overlay. One 600µm2 box containing mature intermediate and terminal villi is selected within each quadrant for analysis. **B.** 8x magnification of one box from panel A. Brown anti-CD34 staining highlights fetal vascular endothelium. **C.** Digital pixel analysis of the image in panel B. Red coloring indicates pixels positive for anti-CD34 stain, while blue and yellow colors indicate negative pixels. %FVE (percent fetal vascular endothelial cells) is calculated as the number of positive pixels divided by the total pixels in the four selected boxes combined.

### Statistical Analysis

We summarized variables using means and standard deviations for continuous variables, medians and interquartile ranges for skewed variables, and frequencies and percentages for categorical variables. We used independent t-tests and Cohen’s *d* effect sizes to evaluate crude comparisons between cases and controls for continuous variables, followed by linear regression with covariate adjustment for maternal smoking status during pregnancy (never, quit, or current) and our matching variables[19] of gestational age at delivery and fetal sex for all comparisons and cold ischemia time (i.e., the time between delivery and formalin fixation of placenta tissue) for analyses of %FVE and placental vascular index. Similar analyses were run for categorical variables using Fisher’s exact tests and logistic regression. Finally, we evaluated associations when stratifying by preterm (<37 weeks’ gestational age) versus term delivery.

## Results

### Sample Characteristics

In total, 68 maternal-neonate dyads were included: 34 cases of CHD and 34 gestational age- and fetal sex-matched controls. One dyad from each group was excluded from analysis due to poor tissue staining. Sample characteristics are presented in **Table 1**. We found no differences between cases and controls for maternal age, BMI at the first prenatal visit, self-reported race or ethnicity, marital or insurance status, rate of cesarean birth, or presence of pre-pregnancy hypertension. Mothers of neonates with CHD were significantly more likely to smoke during pregnancy, while mothers of control neonates were more likely to have quit smoking. By design, there were no differences in gestational age at delivery and fetal sex between cases and controls.

**Table 1.**
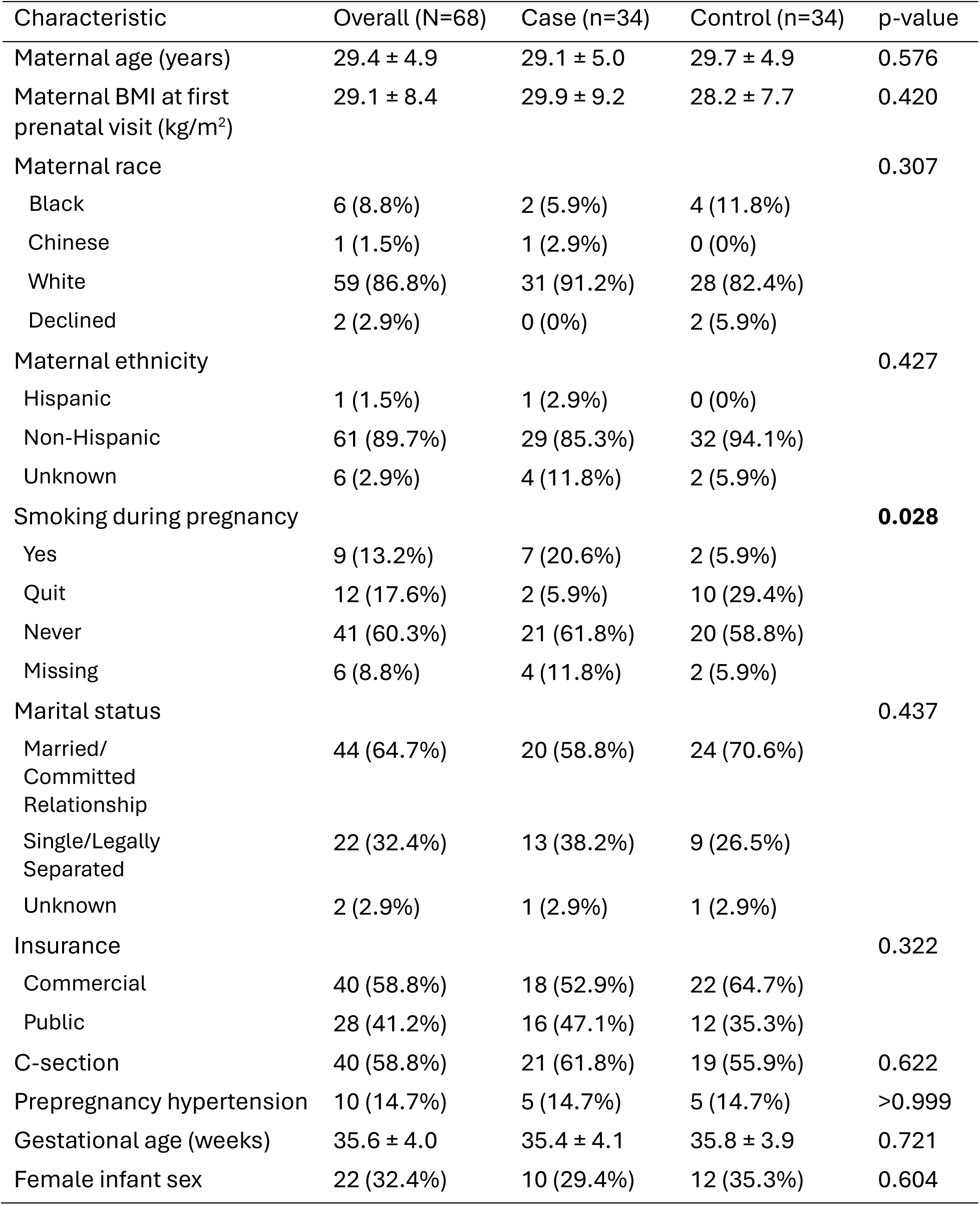
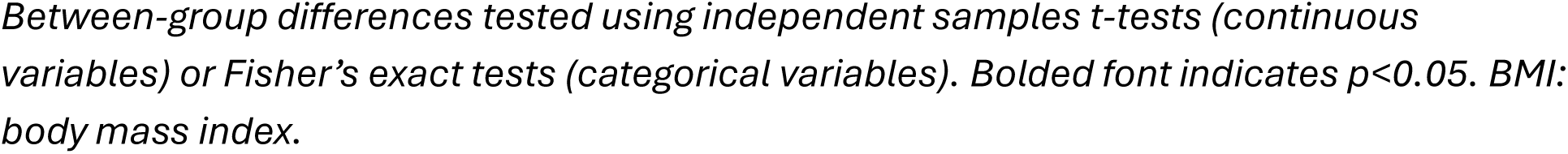
Participant characteristics.

| Characteristic | Overall (N=68) | Case (n=34) | Control (n=34) | p-value |
| --- | --- | --- | --- | --- |
| Maternal age (years) | 29.4 ± 4.9 | 29.1 ± 5.0 | 29.7 ± 4.9 | 0.576 |
| Maternal BMI at first prenatal visit (kg/m<sup>2</sup>) | 29.1 ± 8.4 | 29.9 ± 9.2 | 28.2 ± 7.7 | 0.420 |
| Maternal race |  |  |  | 0.307 |
| Black | 6 (8.8%) | 2 (5.9%) | 4 (11.8%) |  |
| Chinese | 1 (1.5%) | 1 (2.9%) | 0 (0%) |  |
| White | 59 (86.8%) | 31 (91.2%) | 28 (82.4%) |  |
| Declined | 2 (2.9%) | 0 (0%) | 2 (5.9%) |  |
| Maternal ethnicity |  |  |  | 0.427 |
| Hispanic | 1 (1.5%) | 1 (2.9%) | 0 (0%) |  |
| Non-Hispanic | 61 (89.7%) | 29 (85.3%) | 32 (94.1%) |  |
| Unknown | 6 (2.9%) | 4 (11.8%) | 2 (5.9%) |  |
| Smoking during pregnancy |  |  |  | 0.028 |
| Yes | 9 (13.2%) | 7 (20.6%) | 2 (5.9%) |  |
| Quit | 12 (17.6%) | 2 (5.9%) | 10 (29.4%) |  |
| Never | 41 (60.3%) | 21 (61.8%) | 20 (58.8%) |  |
| Missing | 6 (8.8%) | 4 (11.8%) | 2 (5.9%) |  |
| Marital status |  |  |  | 0.437 |
| Married/<br>Committed<br>Relationship | 44 (64.7%) | 20 (58.8%) | 24 (70.6%) |  |
| Single/Legally<br>Separated | 22 (32.4%) | 13 (38.2%) | 9 (26.5%) |  |
| Unknown | 2 (2.9%) | 1 (2.9%) | 1 (2.9%) |  |
| Insurance |  |  |  | 0.322 |
| Commercial | 40 (58.8%) | 18 (52.9%) | 22 (64.7%) |  |
| Public | 28 (41.2%) | 16 (47.1%) | 12 (35.3%) |  |
| C-section | 40 (58.8%) | 21 (61.8%) | 19 (55.9%) | 0.622 |
| Prepregnancy hypertension | 10 (14.7%) | 5 (14.7%) | 5 (14.7%) | >0.999 |
| Gestational age (weeks) | 35.6 ± 4.0 | 35.4 ± 4.1 | 35.8 ± 3.9 | 0.721 |
| Female infant sex | 22 (32.4%) | 10 (29.4%) | 12 (35.3%) | 0.604 |

*Between-group differences tested using independent samples t-tests (continuous variables) or Fisher's exact tests (categorical variables). Bolded font indicates $p < 0.05$ . BMI: body mass index.*

### Placental and Neonatal Outcomes

In the overall sample, we found no statistically significant differences between cases and controls for birthweight, placental weight, placental efficiency, %FVE, or placental vascular index, though unadjusted Cohen’s *d* effect sizes ranged from small (0.20) to moderate (0.41) in the expected directions (**Supplemental Table 1**). Analyses stratified by preterm birth status are presented in **Table 2**. Among preterm neonates, there was no difference between cases and controls for birthweight (Cohen’s *d* effect size [95% confidence interval]: −0.09 [−0.75, 0.58]) but small-to-moderate effect sizes were observed for placental weight (−0.29 [−0.95, 0.38]), placental efficiency (0.39 [−0.28, 1.06]), %FVE (−0.60 [−1.27, 0.09]), and placental vascular index (−0.45 [−1.12, 0.22]). Among term neonates, moderate effect sizes were observed for birthweight (−0.53 [−0.122, 0.17]), placental weight (−0.56 [−1.25, 0.14]), and placental vascular index (−0.39 [−1.07, 0.31]).

**Table 2.**
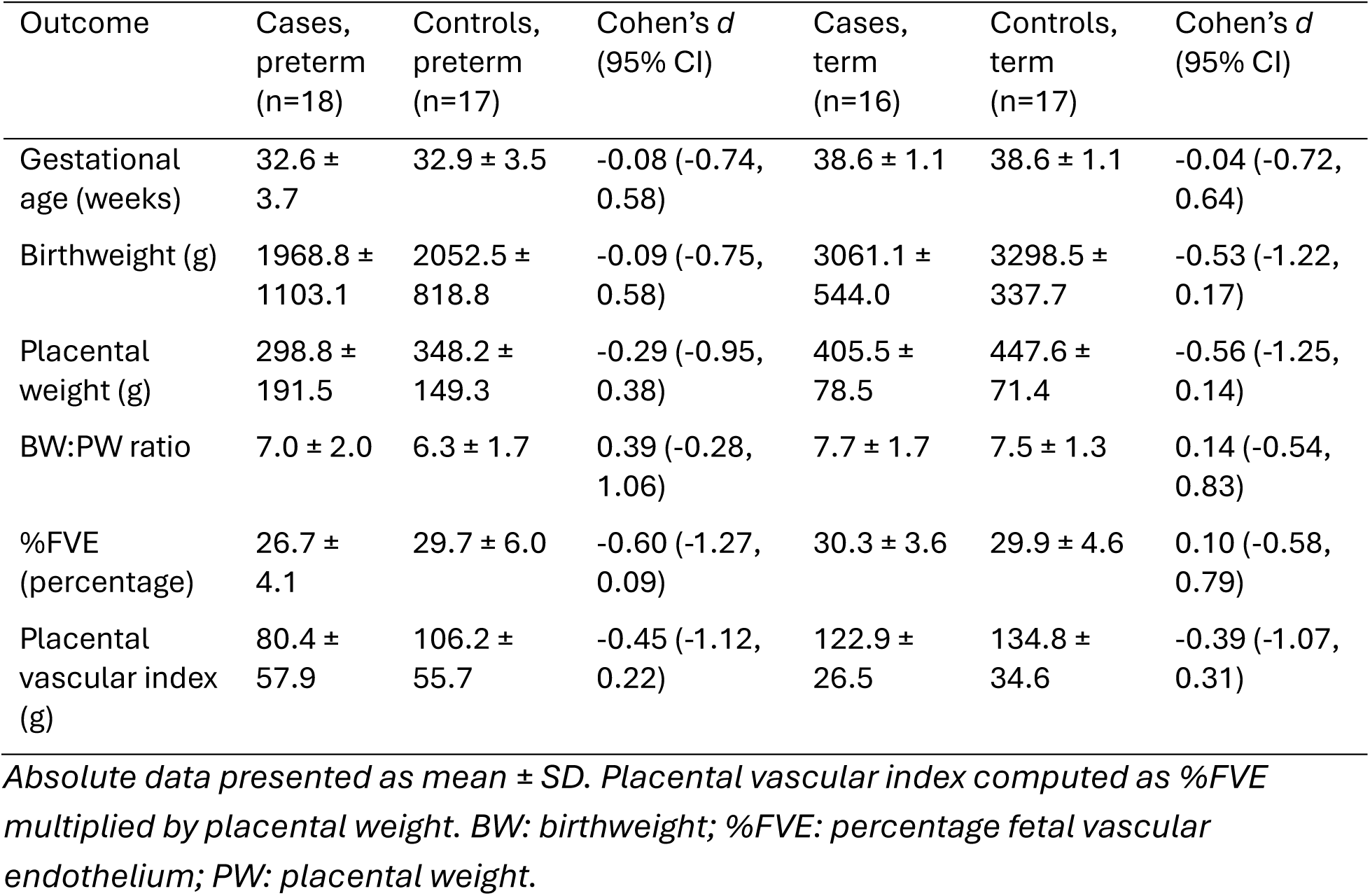
Neonatal birthweight and placental outcomes, stratified by preterm birth status.

| Outcome | Cases,<br>preterm<br>(n=18) | Controls,<br>preterm<br>(n=17) | Cohen's *d*<br>(95% CI) | Cases,<br>term<br>(n=16) | Controls,<br>term<br>(n=17) | Cohen's *d*<br>(95% CI) |
| --- | --- | --- | --- | --- | --- | --- |
| Gestational<br>age (weeks) | 32.6 ±<br>3.7 | 32.9 ± 3.5 | -0.08 (-0.74,<br>0.58) | 38.6 ± 1.1 | 38.6 ± 1.1 | -0.04 (-0.72,<br>0.64) |
| Birthweight (g) | 1968.8 ±<br>1103.1 | 2052.5 ±<br>818.8 | -0.09 (-0.75,<br>0.58) | 3061.1 ±<br>544.0 | 3298.5 ±<br>337.7 | -0.53 (-1.22,<br>0.17) |
| Placental<br>weight (g) | 298.8 ±<br>191.5 | 348.2 ±<br>149.3 | -0.29 (-0.95,<br>0.38) | 405.5 ±<br>78.5 | 447.6 ±<br>71.4 | -0.56 (-1.25,<br>0.14) |
| BW:PW ratio | 7.0 ± 2.0 | 6.3 ± 1.7 | 0.39 (-0.28,<br>1.06) | 7.7 ± 1.7 | 7.5 ± 1.3 | 0.14 (-0.54,<br>0.83) |
| %FVE<br>(percentage) | 26.7 ±<br>4.1 | 29.7 ± 6.0 | -0.60 (-1.27,<br>0.09) | 30.3 ± 3.6 | 29.9 ± 4.6 | 0.10 (-0.58,<br>0.79) |
| Placental<br>vascular index<br>(g) | 80.4 ±<br>57.9 | 106.2 ±<br>55.7 | -0.45 (-1.12,<br>0.22) | 122.9 ±<br>26.5 | 134.8 ±<br>34.6 | -0.39 (-1.07,<br>0.31) |

*Absolute data presented as mean ± SD. Placental vascular index computed as %FVE multiplied by placental weight. BW: birthweight; %FVE: percentage fetal vascular endothelium; PW: placental weight.*

Stratified regression analyses controlling for smoking status (all analyses) and cold ischemia time (%FVE and placental vascular index analyses only) did not detect significant differences. However, notable differences in the magnitude of associations were observed when stratifying by preterm birth status (**Table 3**). While birthweight effect sizes of differences between cases and controls were smaller in both preterm and term CHD neonates, differences were more pronounced among term deliveries. Placental parameters demonstrated the opposite trend, with larger effect sizes of differences between cases and controls observed among preterm neonates (**Figure 2**). A complete breakdown of neonatal and placental parameters by congenital heart disease diagnosis is presented in **Supplemental Table 2**. Finally, the odds of delivering a small-for-gestational-age neonate were 3.7 times higher (95% CI 0.91, 15.22, p=0.068) for cases compared to controls in the overall sample (**Supplemental Table 3**).

**Figure 2.**
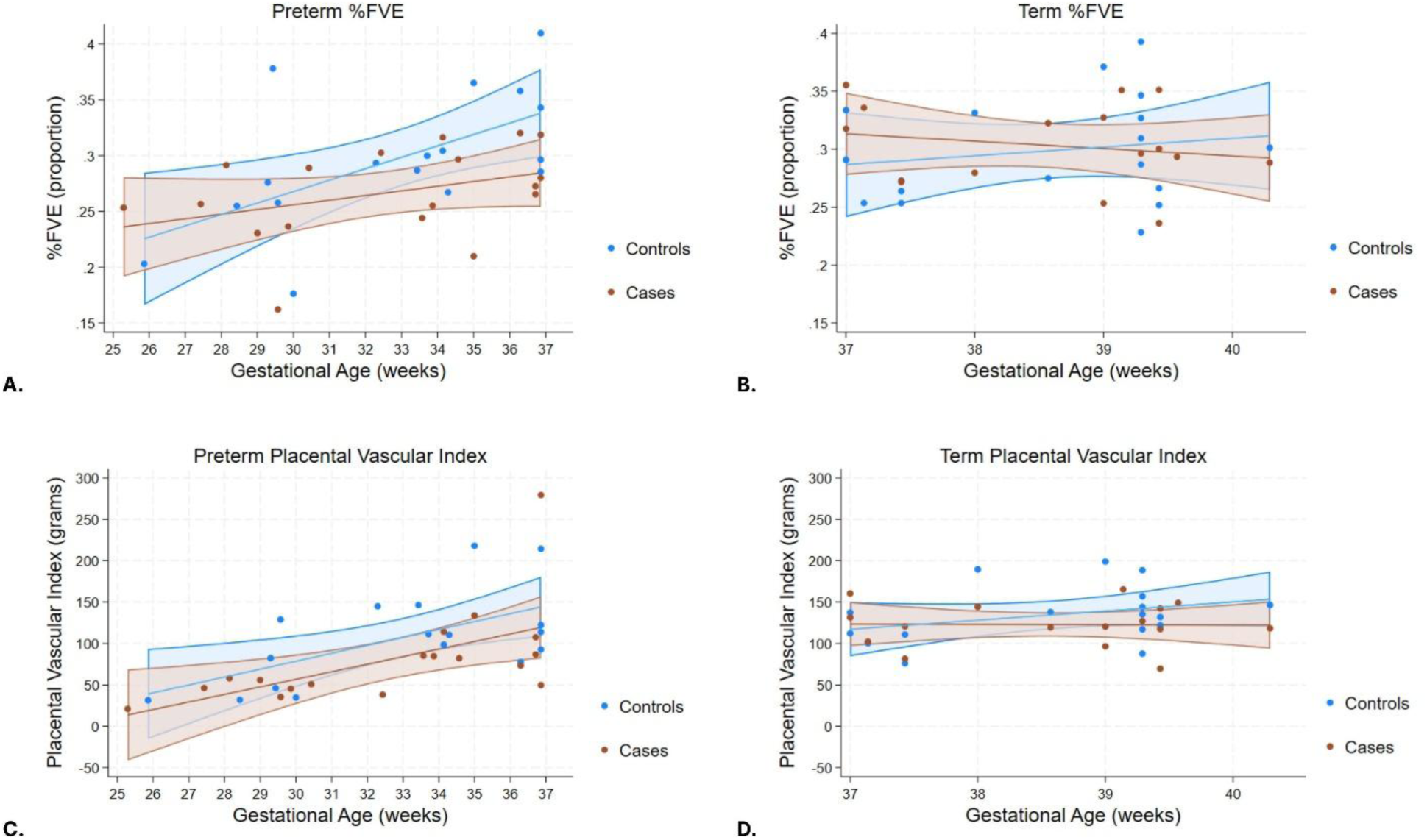
Scatter plots with 95% confidence bands for %FVE (percent fetal vascular endothelial cells, panels A & B) and placental vascular index (panels C & D) among preterm (panels A & C) and term (panels B & D) neonates, plotted against gestational age. Placental vascular index is calculated as placental weight multiplied by %FVE.

**Table 3.** Association of cases compared to controls with neonatal outcomes, stratified by preterm birth status.

| Outcome | Among preterm neonates<br>[$\beta$ coefficient (95% CI)] | p-value | Among term neonates<br>[$\beta$ coefficient (95% CI)] | p-value |
| --- | --- | --- | --- | --- |
| Birthweight (g) | -196.4 (-769.4, 376.6) | 0.488 | -239.3 (-536.4, 57.8) | 0.110 |
| Placental weight (g) | -66.6 (-183.5, 50.4) | 0.253 | -41.2 (-100.9, 18.5) | 0.167 |
| Placental efficiency | 0.84 (-0.66, 2.34) | 0.259 | 0.24 (-1.02, 1.51) | 0.698 |
| %FVE (percentage) | -1.5 (-4.7, 1.6) | 0.332 | -0.001 (-3.1, 3.1) | 0.995 |
| Placental vascular index (g) | -25.7 (-64.9, 13.4) | 0.189 | -13.0 (-37.3, 11.2) | 0.277 |

*$\beta$ corresponds to the difference in outcome in cases compared to controls, adjusted for smoking status, gestational age, and fetal sex (all models) and cold ischemia time (%FVE and placental vascular index only). Preterm: less than 37 completed weeks of gestation; Term: 37 or greater completed weeks of gestation. %FVE: percentage fetal vascular endothelium*

### Maternal Outcomes

Incidence of hypertensive disorders of pregnancy and gestational diabetes were not significantly different between cases and controls (**Table 4**). A higher proportion of women in the control group developed gestational hypertension (cases: 6% vs. controls: 12%) and severe preeclampsia (6% vs. 18%), while a higher proportion of women in the cases group developed preeclampsia superimposed on pre-pregnancy hypertension (15% vs. 3%).

**Table 4.** Maternal outcomes by group.

| Outcome | Overall (N=68) | Case (n=34) | Control (n=34) | p-value |
| --- | --- | --- | --- | --- |
| Hypertensive Disorders of Pregnancy |  |  |  | 0.175 |
| None | 47 (69.1%) | 24 (70.6%) | 23 (67.7%) |  |
| Gestational hypertension | 6 (8.8%) | 2 (5.9%) | 4 (11.8%) |  |
| Mild preeclampsia | 1 (1.5%) | 1 (2.9%) | 0 (0%) |  |
| Severe preeclampsia/HELLP | 8 (11.8%) | 2 (5.9%) | 6 (17.6%) |  |
| Eclampsia | 0 (0%) | 0 (0%) | 0 (0%) |  |
| Preeclampsia superimposed on<br>prepregnancy hypertension | 6 (8.8%) | 5 (14.7%) | 1 (2.9%) |  |
| Diabetes |  |  |  | >0.999 |
| No diabetes | 61 (89.7%) | 30 (88.2%) | 31 (91.2%) |  |
| Gestational DM | 4 (5.9%) | 2 (5.9%) | 2 (5.9%) |  |
| Pregestational T1 or T2 DM | 3 (4.4%) | 2 (5.9%) | 1 (2.9%) |  |

*Data presented as frequency (percentage). Proportions compared between groups using Fisher's exact tests. DM: diabetes mellitus; HELLP: hemolysis, elevated liver enzymes, and low platelets syndrome*

## Discussion

This pilot retrospective case control study found small-to-moderate effect sizes suggesting that neonates diagnosed with CHD may have smaller placentas with lower vascularization of the intermediate and terminal villi. Our findings offer preliminary evidence that CHD neonates born preterm may be subject to a “double hit” of a smaller placenta coupled with lower placental vascularization, but that the proportion of villous tissue occupied by fetal vessels may catch up in neonates that complete at least 37 weeks of gestation. Despite this, lower birthweight among CHD neonates suggests that any accelerated angiogenesis in the final weeks of gestation may not adequately compensate for earlier placental insufficiency.

Placental development is embryologically linked with fetal cardiac development, and it is hypothesized that poor placentation may cause fetal CHD[7]. While preliminary and not reaching statistical significance, our observed effect sizes using this novel vascular quantification technique support prior evidence and suggest potential mechanisms relating to fetal placental angiogenesis. Placentas of fetuses with CHD are more likely to display abnormal growth patterns, including lower weight[15,20,21] and higher proportions affected by delayed villous maturation[15,20,22], chorangiosis[8,15,23], and lower vascular density and area[24,25]. We found that the differences between CHD and control placental parameters were more pronounced in preterm neonates but that the difference in birthweight was more pronounced after 37 weeks of gestation. We speculate that our observation of a lower proportion of villous tissue occupied by fetal vessels prior to 37 weeks of gestation may correspond to delayed villous maturation, while the observed similarity in vascularization between groups at term may suggest that chorangiosis develops later in gestation as a compensatory mechanism to overcome the lower vascularization earlier in development. Future research is required to evaluate these hypotheses.

We also found more pronounced effect sizes for placental efficiency among the preterm subgroup, with preterm CHD placentas trending towards higher efficiency than their control counterparts. Prior evidence is mixed for placental efficiency[15]; some studies find higher efficiency among CHD placentas, while other studies find no difference. Higher efficiency has been alternately described as evidence of an imbalance between fetal and placental growth [26] or of a fetus able to reach its growth potential despite a smaller placenta [15]. However, birthweight and placental weight tend to be lower in the setting of CHD, with higher incidence of weights below the 3rd percentile for each[7,15,21]. In the context of this prior literature, our findings suggest that the ratio of birthweight and placental weight may be too crude to capture insufficient placental vascular contributions to fetal growth.

We did not have large enough numbers of individual CHD diagnoses to evaluate whether placental parameters were more heavily compromised in any particular CHD pathology type. However, as was consistent with our overall analysis, we did find that %FVE was lowest among the diagnoses that most commonly occurred preterm (e.g., atrial septal defect and coarctation of the aorta), and was highest among diagnoses that more commonly occurred after 37 weeks (e.g., transposition of the great arteries). Future research with robust statistical power are required to determine if placental vascular density is different at delivery within different CHD subtypes.

Prior research has found that hypertensive disorders of pregnancy are more common among mothers carrying fetuses affected by CHD[7,14], though we did not replicate this finding in our study. It is possible that due to our gestational age matching criterion, our control dyads were more likely to include women with preterm preeclampsia. In support of this interpretation, a higher proportion of control mothers in our sample developed gestational hypertension or *de novo* preeclampsia but a higher proportion of case mothers developed preeclampsia superimposed on existing pre-pregnancy hypertension. Pre-pregnancy hypertension is a risk factor for CHD, as it likely influences uterine spiral artery remodeling and subsequent placental and fetal cardiac dysfunction[7]. Our results are in line with this prior research.

We were unable to detect significantly different proportions of placental fetal vasculature between groups in our study, which may have been due to women with hypertensive disorders of pregnancy being included in the control group. Our sample of prospectively enrolled dyads with a CHD diagnosis was limited, so we elected to include preterm deliveries to maximize the study sample. However, the inclusion of preterm control dyads meant we did not have a fully “normal” comparator group. It is possible that between-group differences in our placental vascular parameters were muted due to the high proportion of control women with a hypertensive disorder of pregnancy, which is common in preterm birth[27,28]. Future research should evaluate our novel placental vascular quantification metric in those with versus without hypertensive disorders of pregnancy.

### Strengths and Limitations

Our sample was derived from prospectively enrolled participants who agreed to donate tissue samples, which strengthened our analyses by ensuring that we were not reliant on placenta specimens that were collected following referral to a pathologist for clinical reasons. However, by matching on fetal sex and gestational age to minimize effects of these parameters on our placental outcomes we likely biased our control samples towards a higher proportion of preeclampsia outcomes, limiting our ability to detect between-group differences. In addition, we had access to a limited number of participants with more severe forms of CHD; the inclusion of less severe forms may have biased our findings towards the null.

## Conclusions

Our pilot data found small to moderate effect sizes indicating differences in placental parameters between cases with CHD and controls, though differences in our small sample did not achieve statistical significance. We found that, despite higher placental efficiency, placental villous vascularization may be impaired prior to 37 weeks of gestation in the setting of CHD. This suggests that placental efficiency may not capture important differences at the maternal-fetal interface. Further, we found no difference in vascularization in term placentas, suggesting that vascular development may accelerate in the setting of CHD late in pregnancy.

## Data Availability

Data may be available upon reasonable request to the authors.

## Acknowledgements

We gratefully acknowledge the participants who donated placenta tissue for analysis. Funding for this work was provided by an internal award from the Department of Pediatrics at West Virginia University. Data and materials were obtained from the Steve N. Caritis MWRI Magee Obstetrical Maternal Infant (MOMI) Database and Biobank.

## Abbreviations

%FVE: proportion of villous tissue comprised of fetal vascular endothelial cells
CHD: congenital heart disease
MOMI: Magee Obstetric Maternal & Infant

---

**Supplemental Table 1.**
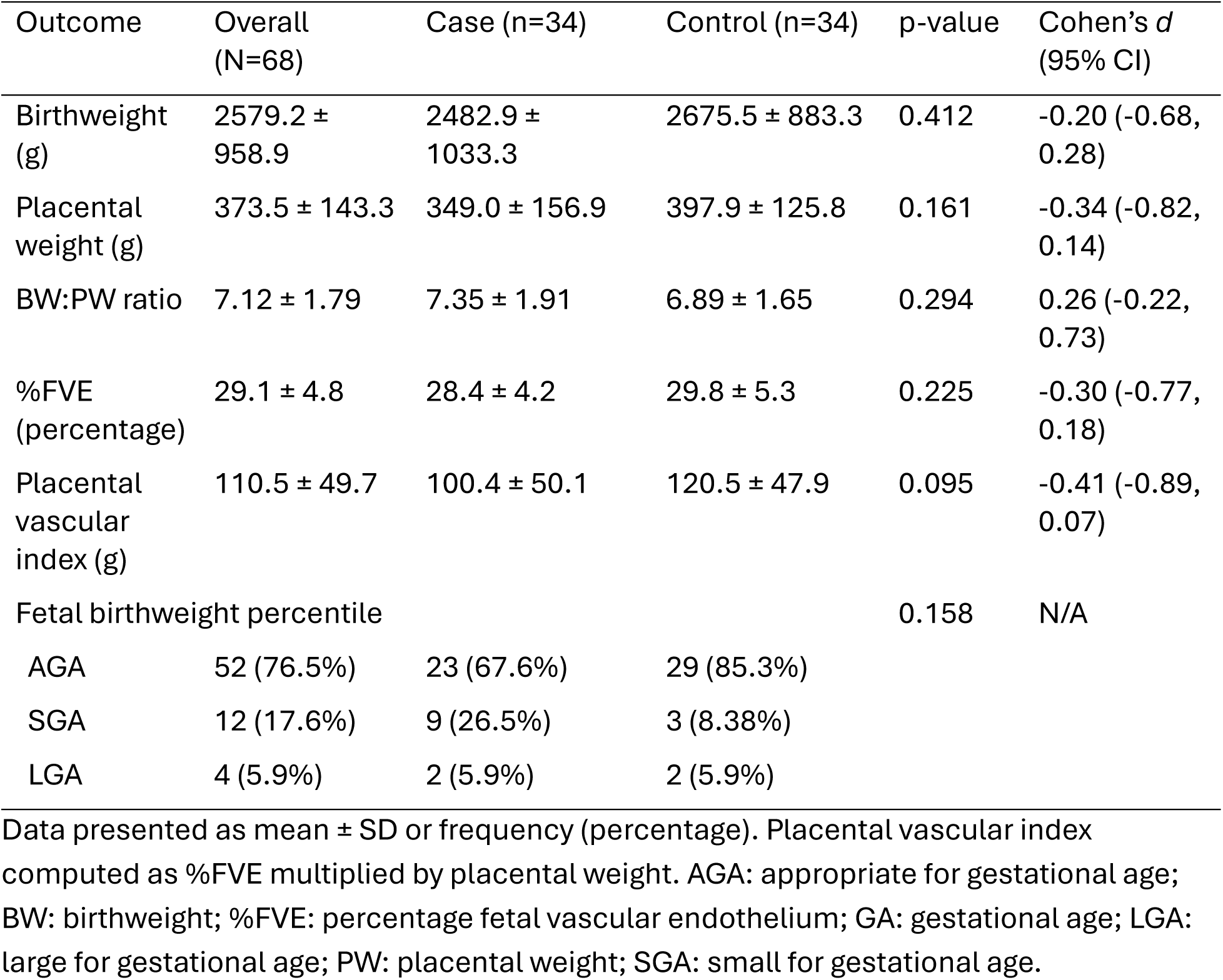
Neonatal birthweight and placental outcomes in the full cohort.

**Supplemental Table 2.** Neonatal birthweight and placental parameters by CHD diagnosis.

| Congenital Heart Defect | n (%) | Birthweight (grams) | Placental weight (grams) | BW:PW ratio | %FVE (percentage) | Placental vascular index (grams) | GA (weeks) |
| --- | --- | --- | --- | --- | --- | --- | --- |
| Atrial Septal Defect | 5 (14.7%) | 1661.6 ± 766.8 | 212.9 ± 108.5 | 8.1 ± 2.7 | 26.2 ± 6.5 | 56.3 ± 35.1 | 31.7 ± 4.5 |
| Ventricular Septal Defect | 9 (26.5%) | 2412 ± 860.6 | 379.1 ± 154.3 | 6.8 ± 2.7 | 28.7 ± 4.0 | 106.3 ± 38.2 | 36.0 ± 3.5 |
| Hypoplastic left heart | 2 (5.9%) | 3430.0 ± 42.4 | 350.0 ± 77.8 | 10.1 ± 2.4 | 29.6 ± 4.4 | 102.0 ± 7.8 | 37.9 ± 1.6 |
| Coarctation of the aorta | 2 (5.9%) | 2315.0 ± 1165.3 | 335.5 ± 132.2 | 6.7 ± 0.8 | 26.3 ± 4.6 | 91.5 ± 50.4 | 34.1 ± 7.3 |
| Severe valve disorders | 2 (5.9%) | 2935.0 ± 275.8 | 380.5 ± 14.8 | 7.7 ± 0.4 | 31.1 ± 1.6 | 118.4 ± 1.3 | 39.0 ± 0.6 |
| Transposition of great arteries | 3 (8.8%) | 3288.7 ± 913.3 | 410.3 ± 95.1 | 8.0 ± 0.6 | 30.6 ± 4.1 | 126.0 ± 34.2 | 38.6 ± 1.6 |
| Tetralogy of Fallot | 4 (11.8%) | 1847.5 ± 567.5 | 279.3 ± 75.1 | 6.6 ± 0.8 | 28.6 ± 2.6 | 81.0 ± 27.8 | 33.4 ± 4.2 |
| Congenital heart block | 2 (5.9%) | 2239.5 ± 425.0 | 350.0 ± 171.1 | 6.9 ± 2.2 | 33.6 ± 2.2 | 119.3 ± 65.0 | 37.7 ± 2.0 |
| Persistent Pulmonary Hypertension | 10 (29.4%) | 2125.5 ± 1017.1 | 264.7 ± 123.6 | 8.1 ± 2.1 | 26.5 ± 5.1 | 71.8 ± 40.1 | 33.6 ± 4.9 |

Data presented as mean ± SD. Placental vascular index computed as %FVE multiplied by placental weight. BW: birthweight; %FVE: percentage fetal vascular endothelium; GA: gestational age; PW: placental weight.

**Supplemental Table 3.**
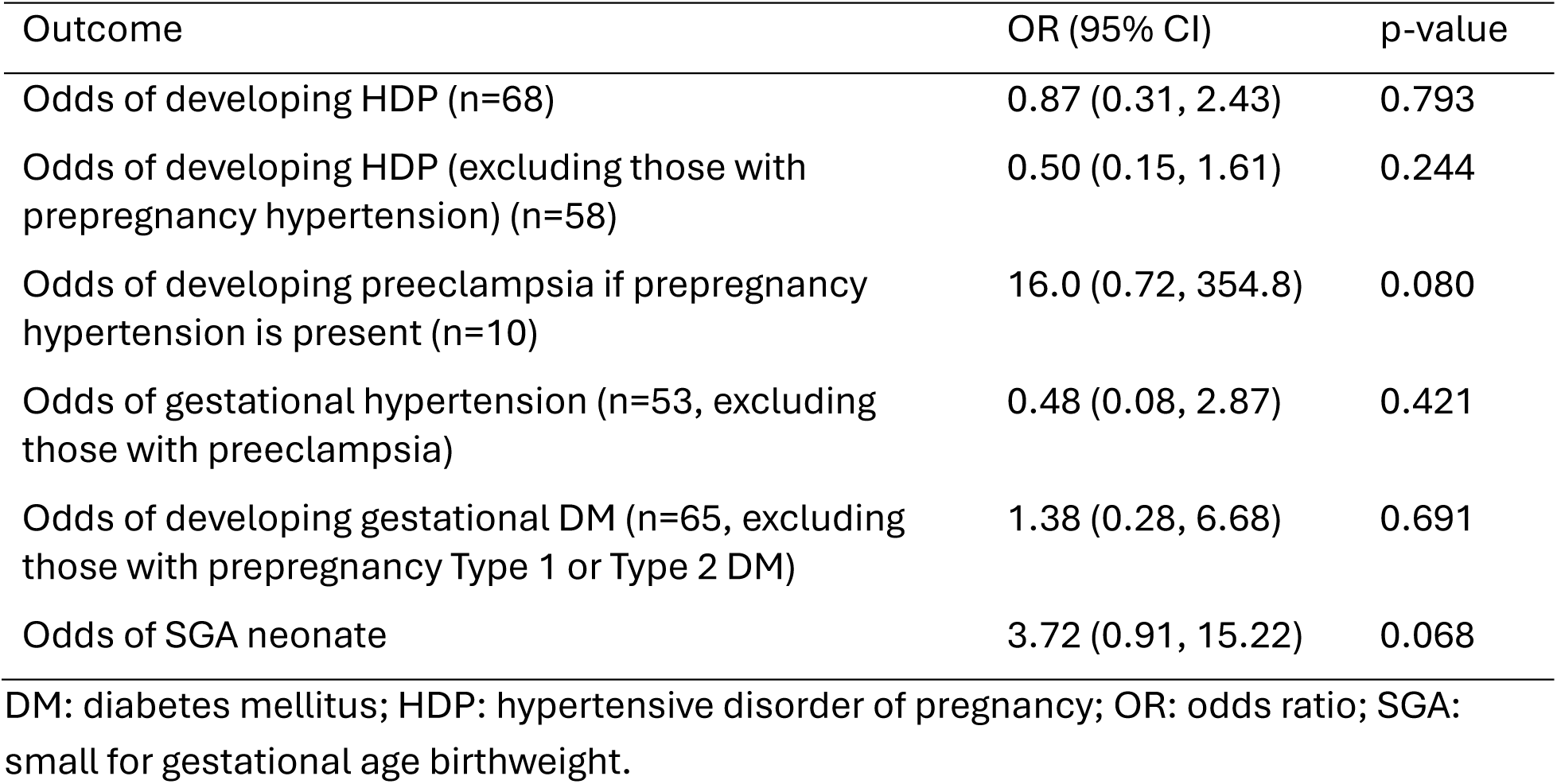
Odds of maternal and fetal outcomes in cases compared to controls.

| Outcome | OR (95% CI) | p-value |
| --- | --- | --- |
| Odds of developing HDP (n=68) | 0.87 (0.31, 2.43) | 0.793 |
| Odds of developing HDP (excluding those with prepregnancy hypertension) (n=58) | 0.50 (0.15, 1.61) | 0.244 |
| Odds of developing preeclampsia if prepregnancy hypertension is present (n=10) | 16.0 (0.72, 354.8) | 0.080 |
| Odds of gestational hypertension (n=53, excluding those with preeclampsia) | 0.48 (0.08, 2.87) | 0.421 |
| Odds of developing gestational DM (n=65, excluding those with prepregnancy Type 1 or Type 2 DM) | 1.38 (0.28, 6.68) | 0.691 |
| Odds of SGA neonate | 3.72 (0.91, 15.22) | 0.068 |

DM: diabetes mellitus; HDP: hypertensive disorder of pregnancy; OR: odds ratio; SGA: small for gestational age birthweight.

